# Long-term exposure to fine particulate matter constituents and incident stroke risk among middle-aged and older adults in China

**DOI:** 10.64898/2025.12.03.25341599

**Authors:** Hao Wang, Miaomiao Wei, Taikun Lu, Chao Wang, Xiaoshuang Xia, Lin Wang, Xin Li

## Abstract

**Background:** While long-term exposure to ambient fine particulate matter (PM_2.5_) is increasingly recognized as a risk factor for stroke, evidence regarding the detrimental effects of its specific constituents remains limited.

**Methods:** We conducted a nationwide prospective cohort study using data from the China Health and Retirement Longitudinal Study (CHARLS) between 2013 and 2018. Individual records were matched with environmental exposure data to quantify the associations between specific PM_2.5_ constituents (sulfate, nitrate, ammonium, organic matter, and black carbon) and the incidence of stroke. We utilized multivariate Cox models to assess the associations between PM_2.5_ constituents and incident stroke, and employed the Quantile-based g-computation model to identify the key contributor among PM_2.5_ constituents.

**Results:** Among 8724 participants enrolled in this study, 610 individuals developed stroke during a median follow-up period of 5.0 years. All five constituents were significantly associated with stroke onset, the adjusted hazard ratios (95% confidence interval [CI]) for each interquartile range increase were 1.55 (95% CI: 1.35-1.79) for PM_2.5_, 1.55 (95% CI: 1.34-1.79) for sulfate, 1.37 (95% CI: 1.18-1.59) for nitrate, 1.42 (95% CI: 1.24-1.63) for ammonium, 1.59 (95% CI: 1.40-1.80) for organic matter, and 1.77 (95% CI: 1.54-2.03) for black carbon, respectively. In the joint effect analysis, black carbon (28.6%) emerged as the primary contributor, followed by organic matter (21.7%) and sulfate (20.4%). Stratified analyses revealed that individuals aged over 65 years and residents of southern China were more susceptible.

**Conclusions:** Long-term exposure to PM_2.5_ constituents is associated with an elevated risk of incident stroke, with black carbon being the primary contributor. Implementing tailored clinical management and targeted regulations on the emissions of carbonaceous components could potentially alleviate the escalating burden of stroke.

## 1 Introduction

Despite significant progress in treatment and prevention, stroke remains the second leading cause of death and a primary source of chronic disability in the world.^1^ In 2021, stroke caused 7.3 million deaths and 160.5 million disability-adjusted life years (DALYs) worldwide.^2^ Pathophysiologically, stroke results from cerebral hemorrhage or obstruction and is clinically categorized into ischemic stroke, intracerebral hemorrhage, and subarachnoid hemorrhage.^3^ Previous compelling evidence indicates that around 90% of stroke cases can be attributed to modifiable risk factors, including air pollution, insufficient physical activity, obesity, unhealthy diet, and smoking.^4,5^ Among these, air pollution is estimated to be responsible for 14% of stroke-related mortality.^6^ Fine particulate matter (PM_2.5_), defined as airborne particles with aerodynamic diameters ≤ 2.5 micrometers, represents a critical environmental pollutant. These microscopic particles demonstrate remarkable permeability through pulmonary alveolar membranes into systemic circulation, potentially inducing neurological damage.^7^

Substantial evidence confirms that both short- and long-term exposure to PM_2.5_ is associated with an increased risk of stroke.^8^ Short-term exposure significantly links to a higher incidence, hospital admission rate, and mortality from stroke, with stronger associations observed for ischemic stroke and among females and the elderly.^9–11^ Beyond these acute health effects, prospective studies in China indicate that long-term PM_2.5_ exposure is associated with an elevated risk of stroke incidence, mortality, and reduced survival, demonstrating robust associations for both ischemic and hemorrhagic subtypes.^12–14^ Apart from its role in primary incidence, a study in the United States revealed that long-term PM_2.5_ exposure adversely affects stroke prognosis by increasing the risk of near-term hospital readmission.^15^ However, the existing literature is not entirely consistent, as some investigations have reported non-significant associations between PM_2.5_ and stroke-related hospitalizations or mortality.^16–18^ This heterogeneity may stem from differences in population characteristics, exposure assessment methods, or variations in the chemical composition of PM_2.5_ across different regions. This emphasizes the significance of pinpointing the specific toxicity of its individual components on stroke.

PM_2.5_ is a complex mixture composed primarily of sulfate, nitrate, ammonium, organic matter, and black carbon, originating from diverse sources like vehicular exhaust, industrial operations, and residential fuel combustion. Each constituent possesses distinct physicochemical and toxicological properties. Consequently, equal increments in PM_2.5_ mass concentration can lead to divergent health effects due to the source-specific composition. This underscores the necessity of evaluating the specific toxicity of a single component to formulate targeted and health-oriented emission control strategies.

Current epidemiological evidence on the associations between specific PM_2.5_ constituents and stroke remains both inconsistent and geographically limited. For instance, a case-crossover study in Boston discovered a connection between elevated black carbon levels and an increased risk of stroke incidence.^19^ Another time-series study in Guangzhou associated all five major PM_2.5_ constituents with stroke mortality.^20^ Nevertheless, findings from these single-city studies may lack generalizability. Moreover, few studies have examined the joint effects of exposure to multiple PM_2.5_ constituents or identified susceptible subpopulations within a nationwide cohort.

To address these knowledge gaps, we conducted a nationwide cohort study in China. Our objectives were as follows: (1) to comprehensively examine the associations between long-term exposure to major PM_2.5_ constituents and first-ever stroke occurrence; (2) to evaluate their joint effects and individual contributions using a novel mixture modeling approach; and (3) to identify potential susceptible subpopulations. We anticipate that our findings will provide a scientific basis for informing more precise and effective emission control strategies for stroke prevention.

## 2 Methods

### 2.1 Study design and population

This prospective cohort study was conducted with China Health and Retirement Longitudinal Study (CHARLS),^21^ an ongoing longitudinal study of middle-aged and older Chinese adults. As a nationally representative survey, CHARLS encompasses participants from 450 communities across 126 cities and counties in 28 provinces in China. Our baseline was established in 2013, coinciding with the implementation of China’s Air Pollution Prevention and Control Action Plan (APPCAP), enabling the examination of associations within a contemporary context of declining air pollution. It is estimated that, compared with the levels in 2013, the annual average concentrations of PM_2.5_ in 2017 witnessed a 33.3% decline in 74 key Chinese cities.^22^ Consequently, 18605 participants were enrolled initially and a total of 10455 participants met the inclusion criteria. At baseline, they had no history of stroke, were 45 years or older, and had complete demographic, health, and physical examination data. Participants were followed up in 2015 and 2018 via face-to-face, computer-assisted personal interviews to track the same survey as in 2013. After excluding 1731 participants due to missing follow-up, 8724 participants were included in the final analysis. The clear participant selection process is presented in Figure S1.

All participants recruited into the CHARLS program were given written informed consent. The CHARLS adheres strictly to the ethical principles outlined in the Declaration of Helsinki and has obtained approval from the Ethics Review Committee of Peking University (IRB00001052–11015).

### 2.2 Assessment of incident stroke

Incident stroke cases were identified based on participants’ responses to specific survey questions regarding a first-ever physician diagnosis during follow-up interviews. This included information from primary care, self-reported data, and hospital admissions. The timing of the stroke event was ascertained by asking participants to report either the year or their age at the time of occurrence. Participants were followed from the 2013 baseline across three interview waves until the time of stroke diagnosis, loss of contact (16.6%), or until the survey end in 2018, whichever occurred first.

### 2.3 Environmental exposure assessment

The daily concentrations of PM_2.5_ and its key constituents, including sulfate, nitrate, ammonium, organic matter and black carbon during the study period were retrieved from Tracking Air Pollution in China (TAP) dataset (http://tapdata.org.cn/). The TAP dataset provides high-resolution estimates by integrating the Weather Research and Forecasting-Community Multiscale Air Quality (WRF-CMAQ) modeling system, ground-based observations, a machine learning-extreme gradient boosting algorithm, and multisource satellite-based PM_2.5_ data, yielding daily predictions at a 10 × 10 km spatial resolution across China.^23,24^ Validation studies have demonstrated strong correlations between TAP estimates and in-situ measurements, with correlation coefficients ranging from 0.67 to 0.80 on a daily scale (2013-2020) and from 0.64 to 0.75 on a monthly scale (2000-2020).^24^ Daily temperature and relative humidity data for the same period were extracted from the European Centre for Medium-Range Weather Forecasts Reanalysis v5 (ERA5) of the global climate.^25^ The ERA5 has a spatial resolution of 0.1° × 0.1° and a temporal resolution of 1 h. To measure environmental exposure, all participants in the CHARLS cohort were geo-coded to the regionalization code of their residential address. The nearest grid cells in the TAP and ERA5 datasets corresponding to the location where the participants were registered were used. The estimates from these grids during the corresponding periods were employed to represent the exposures. The long-term exposure was defined as the average concentration of individuals from the baseline survey until the occurrence of stroke, or the end of the follow-up period.

### 2.4 Covariates

Potential confounders of the associations between pollutant exposure and incident stroke were selected from the CHARLS questionnaire based on previous investigations.^26,27^ The analyses incorporated covariables from sociodemographic characteristics, health-related behaviors, and anthropometric measurements. These covariables encompassed age, gender (female vs. male), education (“primary school or below”, “high school”, and “college or above”), marital status (married vs. unmarried), residence (rural vs. urban), smoking (no vs. yes), drinking (no vs. yes), sleep time, social activity (no vs. yes), hypertension (no vs. yes), dyslipidemia (no vs. yes), diabetes (no vs. yes), heart disease (no vs. yes), systolic blood pressure (SBP), diastolic blood pressure (DBP), body mass index (BMI), waist circumference (WC). All of these variables, in combination with temperature and relative humidity, were employed to exhibit characteristics of participants.

### 2.5 Statistical analysis

Statistical analyses and result visualizations were performed using R software (version 4.4.1). Statistical significance was defined as a two-tailed *p*-value < 0.05. Extreme values in continuous variables were handled by winsorizing at the 1st and 99th percentiles. We compared the characteristics of the participants by stratification of incident stroke. Continuous variables were presented as the mean ± standard deviation (SD), while categorical variables were expressed as frequencies (%). T-tests were employed to analyze continuous variables (such as age, sleep time, and BMI), and chi-square tests were used to examine categorical variables (such as gender, marital status, and hypertension). Pearson correlation analysis was conducted to examine the interrelationships between PM_2.5_ and its constituents. Variance inflation factors (VIF) were calculated to detect multicollinearity, with VIF ≥ 10 indicating serious multicollinearity.

Multivariate Cox models were used to quantify the association between PM_2.5_ constituents and incident stroke. Given the potential multicollinearity among PM_2.5_ constituents, we estimated the hazard ratios (HRs) and 95% confidence intervals (CIs) per interquartile range (IQR) increase in each constituent using single-pollutant models. Model 1 did not incorporate any covariates. Model 2 was adjusted for demographic characteristics (age, gender, education, marital status, residence). In model 3, we further adjusted for lifestyle factors (smoking, drinking, sleep time, social activity), clinical conditions (hypertension, dyslipidemia, diabetes, heart disease), anthropometric measures (BMI, WC), and environmental factors (temperature, relative humidity) in addition to the covariates in model 2. To examine possible nonlinear relationships between fine particulate constituents and stroke incidence, exposure-response curves were examined using the restricted cubic spline (RCS) models with three knots at the 10th, 50th, and 90th percentiles. The reference value was set at the median concentration of each pollutant. Analyses were restricted to the 5th-95th percentile range of exposures, and likelihood ratio tests were used to assess nonlinearity. To determine the concentration threshold at which the incident stroke risk began to increase markedly, we applied segmented regression models. The breakpoint in each model, marking the concentration of most significant slope change, was defined as the threshold.

To assess the joint effects of simultaneous exposure to PM_2.5_ constituents, we employed quantile-based g-computation (QGC) to evaluate the overall mixture effect of five PM_2.5_ constituents on incident stroke risk. The QGC model is a parametric statistical method that combines weighted quantile sum (WQS) regression with the flexibility of g-computation.^28^ In our QGC analysis, the five constituents were divided into quartiles (q = 4) to balance model flexibility with statistical power. The QGC model estimated the joint effect of increasing all constituents simultaneously by one quartile, presented as the HR with a 95% CI. Compared to WQS regression, QGC offers several advantages: (1) it relaxes the unidirectional assumption, allowing constituents to have either positive or negative weights; (2) it provides more robust estimation in the presence of multicollinearity; (3) it better adapts to nonlinear exposure-response relationships observed in our preliminary analyses. Component weights were calculated to show the relative contribution of each constituent to the overall mixture effect, with weights summing to 1 in absolute value. The WQS analysis was conducted as a sensitivity analysis using three quantiles (q = 3) and bootstrap resampling (b = 100), maintaining the unidirectional positive effect assumption.

To identify potential susceptible subpopulations, we performed stratified analyses according to age (< 65 vs. ≥ 65 years), gender, education level (primary school or below vs. high school or above), marital status, geographic region (North vs. South China), smoking status, and BMI (< 24 vs. ≥ 24 kg/m²). Educational attainment was divided into two categories to balance the sizes of the groups and ensure statistical robustness. Geographic stratification was based on the Qinling Mountains-Huai River line, a well-recognized boundary that reflects distinct climatic and environmental gradients in China. This regional classification, consistent with prior research,^29^ was derived from participants’ geolocation data. Since this was a post-hoc derived variable that was not collected during the baseline assessment, it was excluded from the baseline characteristics. Interaction terms between each PM_2.5_ constituent and subgroup variables were tested using likelihood ratio tests to assess statistical significance of effect modification.

Comprehensive sensitivity analyses were conducted to test the robustness of our findings: (1) Residual-based toxicity ranking validation: to verify the stability of the identified toxicity hierarchy independent of total PM_2.5_ mass, we calculated residuals for each constituent from linear regression models with total PM_2.5_ mass as the independent variable. These residuals represent constituent-specific variations orthogonal to total mass. We then applied Cox proportional hazards models to these residual values to re-assess the relative toxicity weights; (2) extreme value exclusion: we re-analyzed the associations after excluding extreme exposure values (beyond the 1st and 99th percentiles) for each PM_2.5_ constituent to assess the influence of potential outliers on effect estimates; (3) proportional hazards assumption assessment: we systematically evaluated the proportional hazards assumption using Schoenfeld residuals tests. For constituents violating this assumption (global test *p* < 0.05), we employed stratified Cox models by gender to accommodate time-varying hazard ratios. All sensitivity analyses maintained the same covariate adjustment as in model 3 and expressed effect sizes as HRs per IQR increase to ensure comparability with primary results.

## 3 Results

### 3.1 Characteristics of study participants and exposure distributions

As shown in Table 1, the characteristics were compared between the incident stroke and non-stroke populations. A total of 8724 participants (mean age 59.95 ± 8.86 years; 49.16% male) were included in this prospective cohort study. During follow-up, 610 individuals (7.0%) developed stroke. Compared with the non-stroke group, participants who experienced stroke were significantly older, more likely to be unmarried and current smokers, had shorter sleep duration, and exhibited higher systolic and diastolic blood pressure, BMI, and WC. They also had significantly higher prevalence rates of hypertension (49.18% vs. 24.90%), dyslipidemia (21.80% vs. 10.67%), diabetes (12.95% vs. 6.51%), and heart disease (23.44% vs. 12.24%). In terms of environmental exposure, stroke cases were associated with significantly lower ambient temperature and relative humidity. The average concentrations of PM_2.5_ total mass, sulfate, nitrate, ammonium, organic matter and black carbon during the study period were 50.77, 9.34, 10.77, 7.29, 12.35, and 2.43 μg/m^3^, respectively (Table 2). The spatial distributions of ambient PM_2.5_ and its five constituents across China are presented in Figure S2. Strong positive correlations were observed between PM_2.5_ and its constituents, as well as among the constituents themselves (Spearman correlation coefficient r > 0.95; Figure S3). Consequently, VIF analysis revealed severe multicollinearity, with all VIFs substantially exceeding 10 (Figure S4).

**Table 1.**
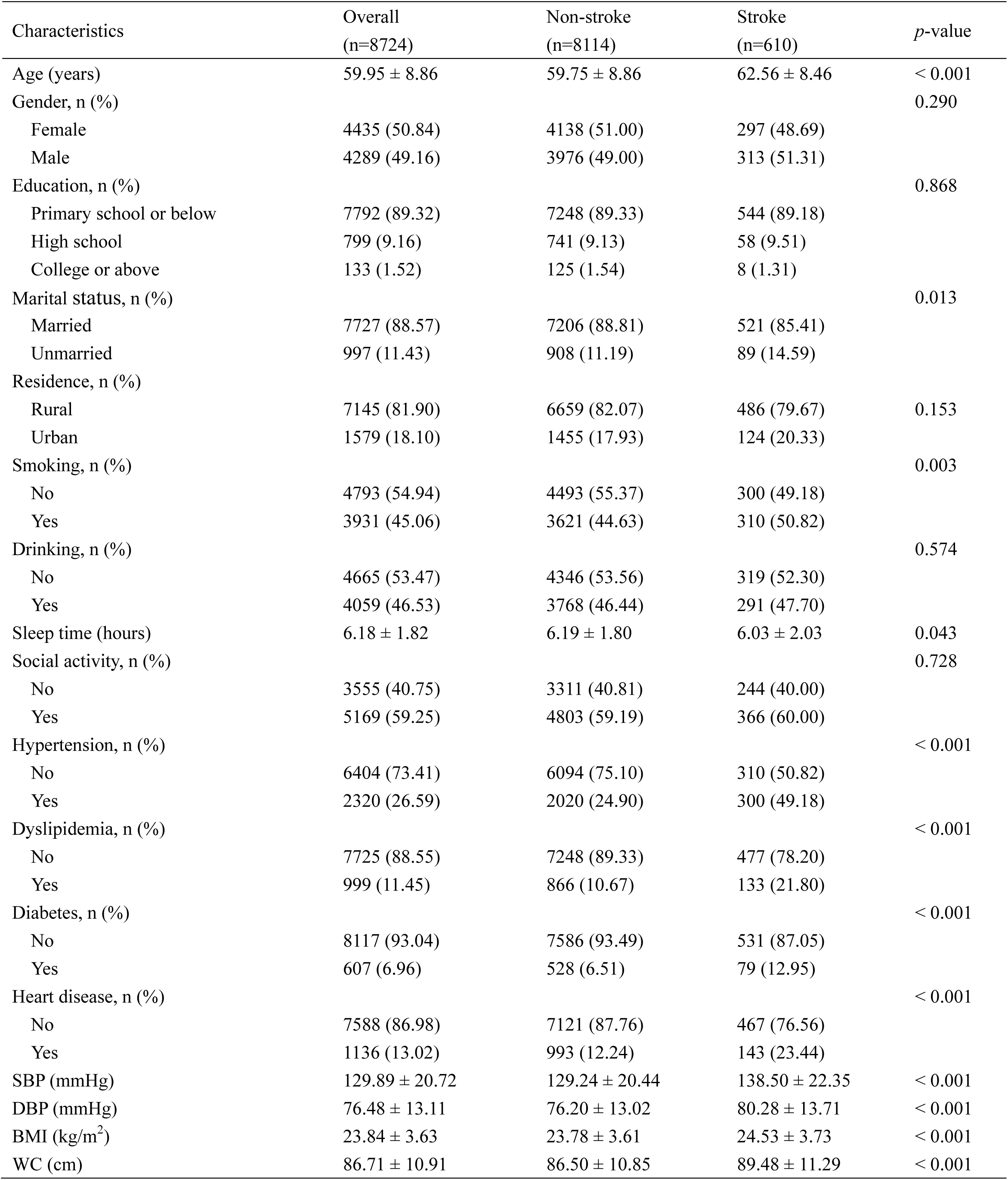
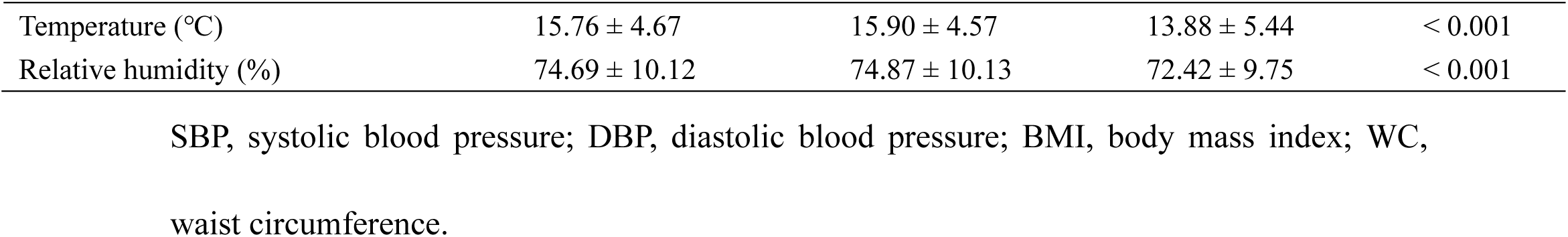
The characteristics of participants by incident stroke status during follow-up.

| Characteristics | Overall<br>(n=8724) | Non-stroke<br>(n=8114) | Stroke<br>(n=610) | *p*-value |
| --- | --- | --- | --- | --- |
| Age (years) | 59.95 ± 8.86 | 59.75 ± 8.86 | 62.56 ± 8.46 | < 0.001 |
| Gender, n (%) |  |  |  | 0.290 |
| Female | 4435 (50.84) | 4138 (51.00) | 297 (48.69) |  |
| Male | 4289 (49.16) | 3976 (49.00) | 313 (51.31) |  |
| Education, n (%) |  |  |  | 0.868 |
| Primary school or below | 7792 (89.32) | 7248 (89.33) | 544 (89.18) |  |
| High school | 799 (9.16) | 741 (9.13) | 58 (9.51) |  |
| College or above | 133 (1.52) | 125 (1.54) | 8 (1.31) |  |
| Marital status, n (%) |  |  |  | 0.013 |
| Married | 7727 (88.57) | 7206 (88.81) | 521 (85.41) |  |
| Unmarried | 997 (11.43) | 908 (11.19) | 89 (14.59) |  |
| Residence, n (%) |  |  |  |  |
| Rural | 7145 (81.90) | 6659 (82.07) | 486 (79.67) | 0.153 |
| Urban | 1579 (18.10) | 1455 (17.93) | 124 (20.33) |  |
| Smoking, n (%) |  |  |  | 0.003 |
| No | 4793 (54.94) | 4493 (55.37) | 300 (49.18) |  |
| Yes | 3931 (45.06) | 3621 (44.63) | 310 (50.82) |  |
| Drinking, n (%) |  |  |  | 0.574 |
| No | 4665 (53.47) | 4346 (53.56) | 319 (52.30) |  |
| Yes | 4059 (46.53) | 3768 (46.44) | 291 (47.70) |  |
| Sleep time (hours) | 6.18 ± 1.82 | 6.19 ± 1.80 | 6.03 ± 2.03 | 0.043 |
| Social activity, n (%) |  |  |  | 0.728 |
| No | 3555 (40.75) | 3311 (40.81) | 244 (40.00) |  |
| Yes | 5169 (59.25) | 4803 (59.19) | 366 (60.00) |  |
| Hypertension, n (%) |  |  |  | < 0.001 |
| No | 6404 (73.41) | 6094 (75.10) | 310 (50.82) |  |
| Yes | 2320 (26.59) | 2020 (24.90) | 300 (49.18) |  |
| Dyslipidemia, n (%) |  |  |  | < 0.001 |
| No | 7725 (88.55) | 7248 (89.33) | 477 (78.20) |  |
| Yes | 999 (11.45) | 866 (10.67) | 133 (21.80) |  |
| Diabetes, n (%) |  |  |  | < 0.001 |
| No | 8117 (93.04) | 7586 (93.49) | 531 (87.05) |  |
| Yes | 607 (6.96) | 528 (6.51) | 79 (12.95) |  |
| Heart disease, n (%) |  |  |  | < 0.001 |
| No | 7588 (86.98) | 7121 (87.76) | 467 (76.56) |  |
| Yes | 1136 (13.02) | 993 (12.24) | 143 (23.44) |  |
| SBP (mmHg) | 129.89 ± 20.72 | 129.24 ± 20.44 | 138.50 ± 22.35 | < 0.001 |
| DBP (mmHg) | 76.48 ± 13.11 | 76.20 ± 13.02 | 80.28 ± 13.71 | < 0.001 |
| BMI (kg/m<sup>2</sup>) | 23.84 ± 3.63 | 23.78 ± 3.61 | 24.53 ± 3.73 | < 0.001 |
| WC (cm) | 86.71 ± 10.91 | 86.50 ± 10.85 | 89.48 ± 11.29 | < 0.001 |
| Temperature (°C) | 15.76 ± 4.67 | 15.90 ± 4.57 | 13.88 ± 5.44 | < 0.001 |
| Relative humidity (%) | 74.69 ± 10.12 | 74.87 ± 10.13 | 72.42 ± 9.75 | < 0.001 |

SBP, systolic blood pressure; DBP, diastolic blood pressure; BMI, body mass index; WC, waist circumference.

**Table 2.** The distributions of PM_2.5_ and its constituents during the study period.

| Variables | Mean | SD | Minimum | P<sub>25</sub> | Median | P<sub>75</sub> | Maximum |
| --- | --- | --- | --- | --- | --- | --- | --- |
| PM<sub>2.5</sub> (µg/m<sup>3</sup>) | 50.77 | 18.32 | 17.15 | 35.58 | 49.10 | 63.32 | 118.13 |
| Sulfate (µg/m<sup>3</sup>) | 9.34 | 3.01 | 3.40 | 6.65 | 9.29 | 11.78 | 19.66 |
| Nitrate (µg/m<sup>3</sup>) | 10.77 | 4.85 | 2.48 | 6.66 | 10.96 | 14.93 | 26.51 |
| Ammonium (µg/m<sup>3</sup>) | 7.29 | 2.92 | 2.25 | 4.81 | 7.51 | 9.53 | 16.72 |
| Organic matter (µg/m<sup>3</sup>) | 12.35 | 4.09 | 4.46 | 9.15 | 11.83 | 14.78 | 35.70 |
| Black carbon (µg/m<sup>3</sup>) | 2.43 | 0.67 | 1.10 | 1.94 | 2.33 | 2.94 | 5.67 |

SD, standard deviation; P<sub>25</sub>, 25th percentile (first quartile); P<sub>75</sub>, 75th percentile (third quartile);
PM<sub>2.5</sub>, particulate matter with aerodynamic diameter $\leq 2.5$ µm.

### 3.2 Associations of PM_2.5_ constituents with stroke incidence

During a median follow-up of 5.0 years (range: 0.17-5.25 years), corresponding to 42354 person-years of observation, the stroke incidence rate was 14.4 per 1000 person-years. In the single-pollutant Cox models, each IQR increase in PM_2.5_ and its constituents was significantly associated with a higher risk of incident stroke across all three models, with all *p*-values < 0.001 (Table 3). In the fully adjusted model (model 3), the HRs were highest for black carbon (HR = 1.77, 95% CI: 1.54-2.03), followed by organic matter (HR = 1.59, 95% CI: 1.40-1.80), PM_2.5_ total mass (HR = 1.55, 95% CI: 1.35-1.79), sulfate (HR = 1.55, 95% CI: 1.34-1.79), ammonium (HR = 1.42, 95% CI: 1.24-1.63), and nitrate (HR = 1.37, 95% CI: 1.18-1.59). The exposure-response relationships of PM_2.5_ and its constituents with stroke incidence were all significantly non-linear (all *p* for nonlinearity < 0.001), exhibiting U-shaped patterns as depicted in Figure 1. Detailed metrics of these curves are provided in Table S1. Notably, black carbon demonstrated the most pronounced health risk, characterized by the highest maximum HR (16.75), the lowest concentration threshold (3.5 μg/m^3^), and the largest area under the curve (AUC).

**Figure 1.**
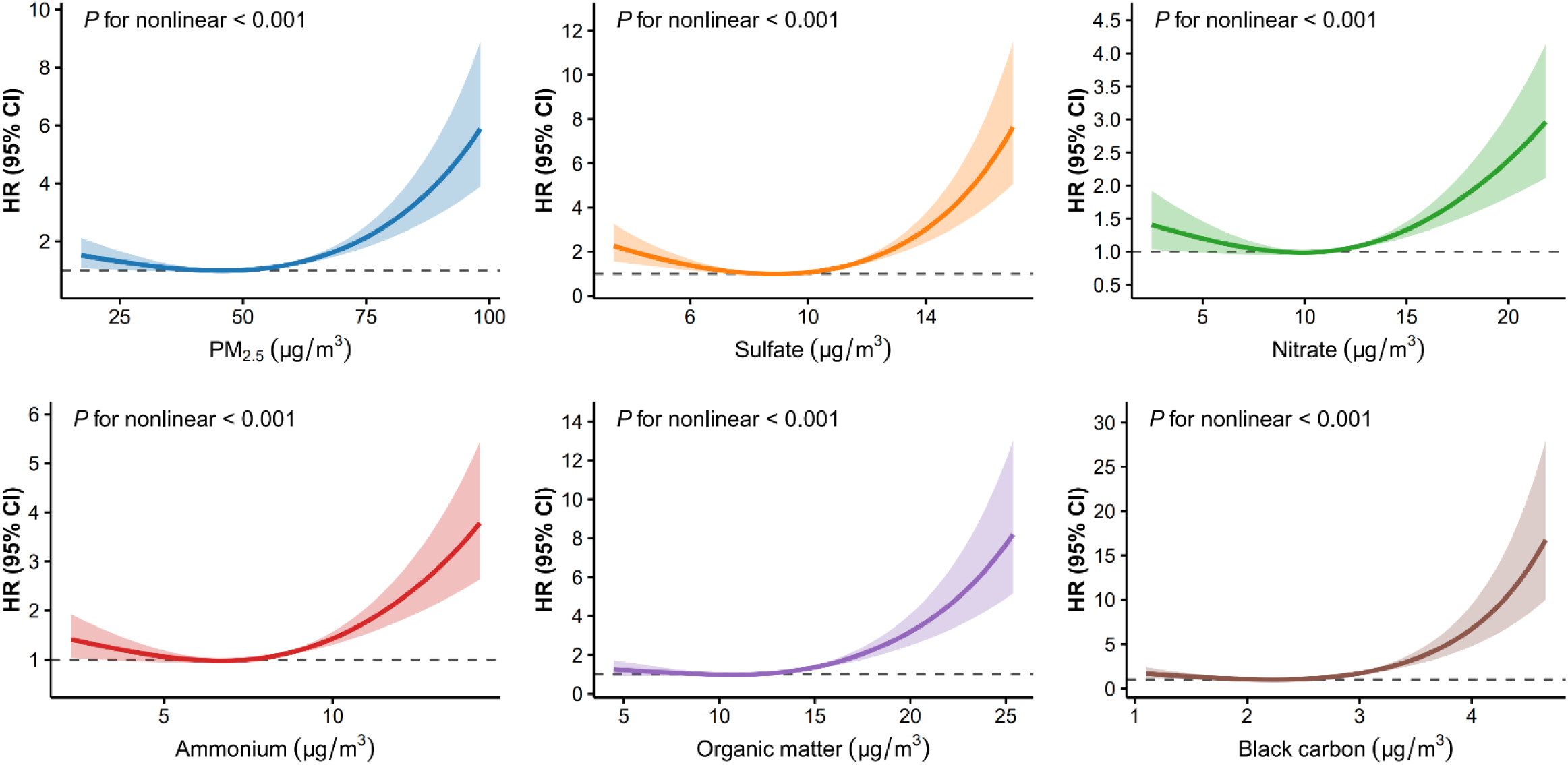
Exposure-response relationships of PM_2.5_ and its constituents with the risk of incident stroke. The model was adjusted for age, gender, education, marital status, residence, smoking, drinking, sleep time, social activity, hypertension, dyslipidemia, diabetes, heart disease, body mass index, waist circumference, temperature, and relative humidity. HR, hazard ratio; CI, confidence interval; PM_2.5_, particulate matter with aerodynamic diameter ≤ 2.5 μm.

**Table 3.** Association of PM_2.5_ and its constituents with the risk of incident stroke by the single-pollutant model.

| Pollutants | IQR | Model 1 <sup>*</sup><br>HR (95% CI) | *p*-value | Model 2 <sup>†</sup><br>HR (95% CI) | *p*-value | Model 3 <sup>‡</sup><br>HR (95% CI) | *p*-value |
| --- | --- | --- | --- | --- | --- | --- | --- |
| PM<sub>2.5</sub> | 27.73 | 1.49 (1.32-1.68) | < 0.001 | 1.50 (1.33-1.69) | < 0.001 | 1.55 (1.35-1.79) | < 0.001 |
| Sulfate | 5.13 | 1.48 (1.29-1.69) | < 0.001 | 1.47 (1.28-1.69) | < 0.001 | 1.55 (1.34-1.79) | < 0.001 |
| Nitrate | 8.27 | 1.43 (1.25-1.64) | < 0.001 | 1.43 (1.25-1.64) | < 0.001 | 1.37 (1.18-1.59) | < 0.001 |
| Ammonium | 4.72 | 1.43 (1.26-1.63) | < 0.001 | 1.43 (1.26-1.63) | < 0.001 | 1.42 (1.24-1.63) | < 0.001 |
| Organic matter | 5.63 | 1.44 (1.30-1.60) | < 0.001 | 1.45 (1.30-1.61) | < 0.001 | 1.59 (1.40-1.80) | < 0.001 |
| Black carbon | 1.00 | 1.54 (1.37-1.73) | < 0.001 | 1.55 (1.38-1.74) | < 0.001 | 1.77 (1.54-2.03) | < 0.001 |

IQR, interquartile range; HR, hazard ratio; CI, confidence interval; PM<sub>2.5</sub>, particulate matter with aerodynamic diameter $\leq 2.5$ $\mu\text{m}$ .
<sup>\*</sup> Model 1 did not incorporate any covariates.
<sup>†</sup> Model 2 was adjusted for age, gender, education, marital status and residence.
<sup>‡</sup> Model 3 was adjusted for age, gender, education, marital status, residence, smoking, drinking, sleep time, social activity, hypertension, dyslipidemia, diabetes, heart disease, body mass index, waist circumference, temperature, and relative humidity.

### 3.3 Joint effects of PM_2.5_ constituent mixtures

The results from the QGC analysis are presented in Figure 2. We observed a significant positive association between the joint exposure to the PM_2.5_ mixture and incident stroke. Each one-quantile increase in the overall mixture was associated with a 20.90% higher risk of stroke (HR = 1.21, 95% CI: 1.11-1.31). When scaled per IQR increase, the risk was substantially elevated by 46.10% (HR = 1.46, 95% CI: 1.24-1.72). Among all constituents, black carbon was identified as the primary contributor, accounting for the largest weight (28.61%), followed by organic matter (21.70%) and sulfate (20.40%).

**Figure 2.**
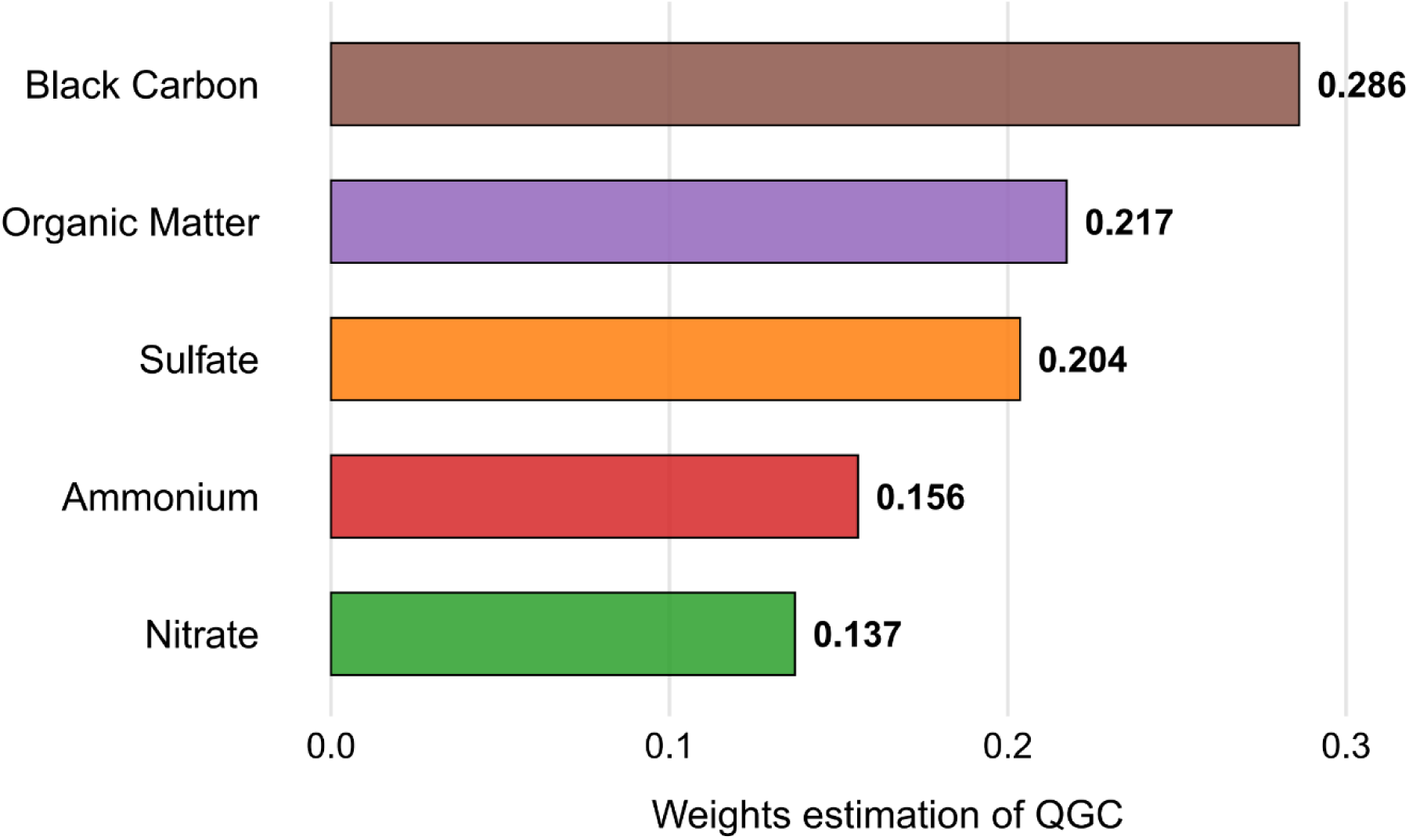
Weights of PM_2.5_ constituents in the mixture index for the association between the particle mixture and incident stroke by QGC. The model was adjusted for age, gender, education, marital status, residence, smoking, drinking, sleep time, social activity, hypertension, dyslipidemia, diabetes, heart disease, body mass index, waist circumference, temperature, and relative humidity. PM_2.5_, particulate matter with aerodynamic diameter ≤ 2.5 μm; QGC, quantile-based g-computation.

### 3.4 Stratified analyses

Stratified analyses revealed significant effect modifications by age and geographic region on the associations between PM_2.5_ and its constituents with incident stroke risk (Figure 3). The adverse effects of PM_2.5_ and its constituents were consistently stronger among older adults (≥ 65 years) compared to their younger counterparts (< 65 years). These differences were statistically significant for black carbon (*p* for interaction = 0.002), organic matter (*p* for interaction = 0.006), sulfate (*p* for interaction = 0.017), and ammonium (*p* for interaction = 0.048). A pronounced North-South geographic disparity was also observed. Residents of Southern China exhibited significantly heightened vulnerability to all pollutants assessed (all *p* for interaction < 0.001), with particularly strong associations for black carbon (HR = 3.00, 95% CI: 2.17-4.15 in the South vs. HR = 1.18, 95% CI: 1.04-1.34 in the North). In contrast, no statistically significant effect modification was detected for gender, BMI, education, marital status, or smoking status (all *p* for interaction > 0.05).

**Figure 3.**
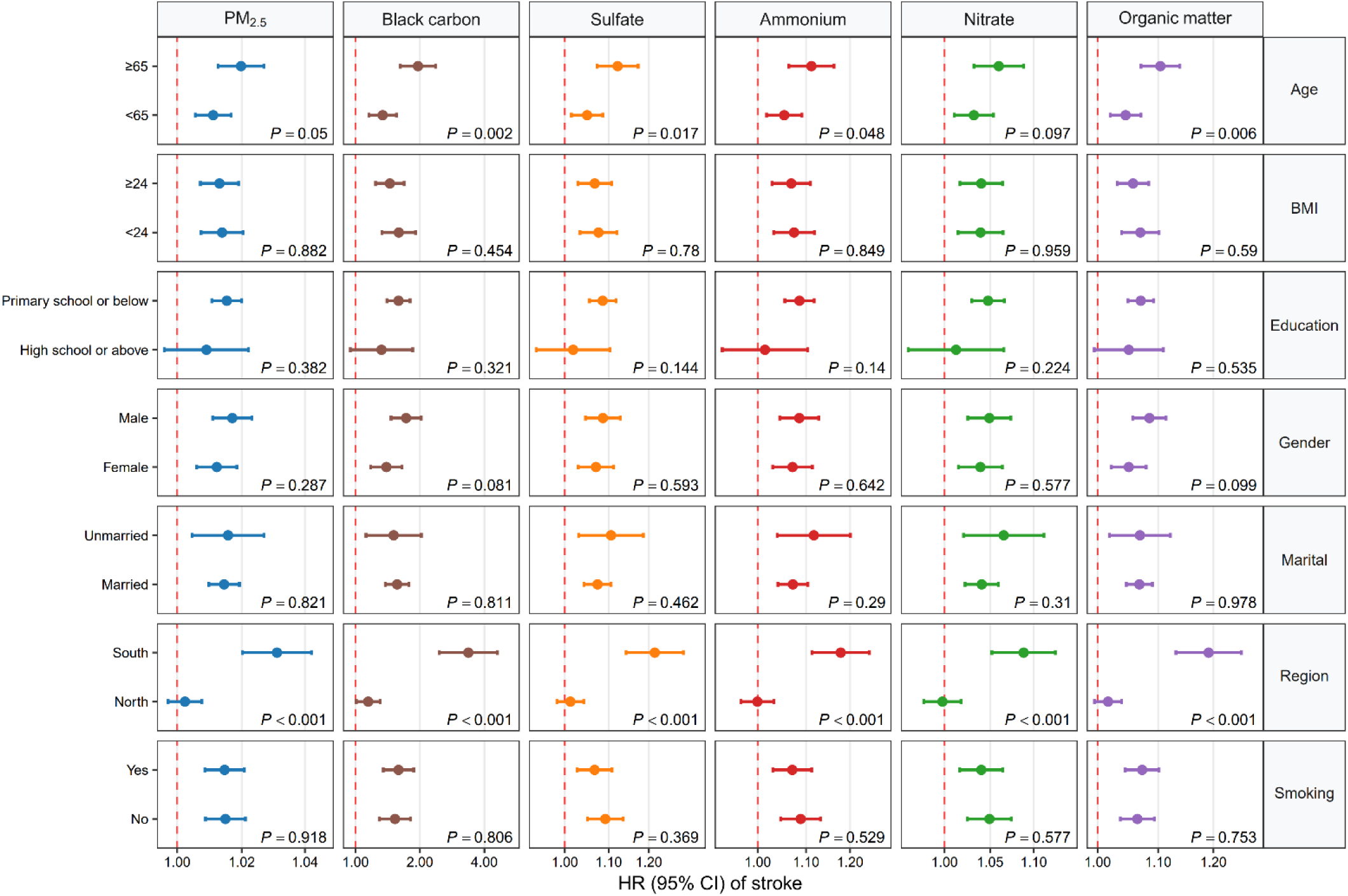
Hazard ratio (HR) (95% confidence interval [CI]) for stroke incidence associated with per interquartile range increase in PM_2.5_ and its constituents, stratified by age, BMI, education, gender, marital status, geographic region, and smoking status. Within each stratum, models were adjusted for all covariates except the stratification variable. The adjustment set included: age, gender, education, marital status, residence, smoking, drinking, sleep time, social activity, hypertension, dyslipidemia, diabetes, heart disease, body mass index, waist circumference, temperature, and relative humidity. The *P* values shown in the figure are for the test of interaction between each pollutant and the stratification variables. PM_2.5_, particulate matter with aerodynamic diameter ≤ 2.5 μm; BMI, body mass index; HR, hazard ratio; CI, confidence interval.

### 3.5 Sensitivity analyses

Sensitivity analyses confirmed the robustness of our primary findings. First, residual-based analysis robustly validated the identified toxicity hierarchy. When examining constituent-specific variations independent of total PM_2.5_ mass, black carbon remained the predominant toxic driver with a HR of 6.84 (95% CI: 3.95-12.15) per IQR increase (1.00 μg/m^3^), accounting for 81.0% of the quantified toxicity. The identical ranking—black carbon > organic matter > sulfate > ammonium > nitrate—between residual-based and original QGC analyses demonstrates the stability of our findings against multicollinearity concerns (Table S2). Second, results were consistent in a WQS regression, which also identified black carbon as the largest contributor to the mixture effect (Weight: 0.192, Figure S5). Furthermore, the associations remained stable across different adjustment models in Cox regression (Figure S6), persisted after excluding extreme exposure values (Table S3), and remained consistent after addressing violations of the proportional hazards assumption (Table S4). Gender-stratified analyses (Table S5) indicated that while associations were generally positive in both genders, the risk estimates appeared higher for black carbon and organic matter in males, and for sulfate, ammonium, and nitrate in females.

## 4 Discussion

This prospective, nationwide cohort study demonstrates that long-term exposure to specific PM_2.5_ constituents (sulfate, nitrate, ammonium, organic matter, and black carbon) is significantly associated with an elevated risk of incident stroke among middle-aged and older Chinese adults. Black carbon was the major contributor to stroke incidence, followed by organic matter and sulfate. These adverse effects were more pronounced in adults over 65 and residents of southern China. To our knowledge, this is the first nationwide prospective cohort study in China to comprehensively assess the single and joint effects of specific PM_2.5_ constituents on incident stroke. Our findings thereby provide a scientific basis for formulating targeted emission control strategies and public health interventions for stroke prevention.

Our findings align with and extend the previous nationwide investigations on the stroke burden attributable to PM_2.5_ constituents. A time-stratified case-crossover study identified positive associations between short-term exposure to specific PM_2.5_ components and ischemic stroke hospital admissions,^30^ with excess risks of 1.77% (95% CI: 0.65-2.90%) for ammonium and 1.69% (95% CI: 0.99-2.39%) for indeno (1, 2, 3-cd) pyrene—a polycyclic aromatic hydrocarbon (PAH) within organic matter. For long-term effects on stroke mortality, a nationwide prospective cohort established significantly increased risks for all five major PM_2.5_ constituents,^31^ with HRs in descending order of magnitude as follows: black carbon (HR = 1.19, 95% CI: 1.07-1.38), nitrate (HR = 1.18, 95% CI: 1.04-1.42), ammonium (HR = 1.18, 95% CI: 1.04-1.41), organic matter (HR = 1.15, 95% CI: 1.05-1.30), and sulfate (HR = 1.14, 95% CI: 1.01-1.34). These results collectively reinforce that multiple PM_2.5_ constituents contribute to stroke risk. Our study significantly advances this field by moving beyond single-pollutant models to dissect the complex mixture effects of PM_2.5_ constituents. The QGC model identified black carbon as the predominant driver, contributing over 28% to the joint effect, followed by organic matter and sulfate. This toxicological hierarchy is strongly corroborated by a large-scale inpatient study on stroke fatality,^32^ which identified the same three components as leading contributors based on population attributable fractions: black carbon (10.6%), organic matter (9.9%), and sulfate (6.6%). The primacy of black carbon was further and strikingly highlighted in our residual-based analysis, where its association with stroke persisted and intensified after accounting for total PM_2.5_ mass. This highlights a critical limitation of regulating PM_2.5_ solely by mass concentration and underscores the imperative to target specific toxic components.

Furthermore, we revealed significant nonlinear exposure-response relationships for all constituents. Whereas the prior inpatient study reported monotonically increasing risk curves,^32^ our analyses demonstrated that the risk of incident stroke increased markedly only after concentrations surpassed specific thresholds. This pattern finds some support in earlier research,^33^ which suggested a potential plateauing of risk at lower concentrations. However, a pivotal finding from our RCS curves is that these thresholds were consistently low, particularly for black carbon (3.5 μg/m^3^), and the risk escalated sharply thereafter. This indicates that there is likely no truly safe level of exposure and that the risk of incident stroke rises appreciably even at concentration ranges considered relatively low. This also implies that moderate reductions in ambient levels of key pollutants, especially black carbon, could confer substantial benefits for primary stroke prevention.

Our findings identified black carbon and organic matter as the two most hazardous constituents, which jointly accounted for over 50% of the quantified incident stroke risk. In China, urban black carbon concentrations typically range from 1.0 to 7.5 μg/m^3^, considerably exceeding levels in most developed regions.^34^ Black carbon originates predominantly from incomplete combustion processes, including vehicular emissions, residential solid fuel burning, and industrial activities.^35^ Toxicologically, black carbon acts as a primary carrier of co-pollutants. Its large specific surface area facilitates the adsorption of various toxic substances, such as PAHs and heavy metals, thereby promoting synergistic effects that exacerbate oxidative stress and systemic inflammation.^36,37^ This is consistent with findings that lead, selenium, and cadmium in PM_2.5_ related to coal combustion were associated with ischemic stroke.^38^ Additionally, black carbon can also directly induce reactive oxygen species (ROS) generation, disrupt cellular calcium homeostasis, and cause mitochondrial dysfunction.^39^ Organic matter, which comprises a complex mixture of numerous compounds, stems from both primary combustion emissions and secondary atmospheric reactions. In addition to driving oxidative and antioxidative imbalances, certain organic matter components, particularly PAHs, are capable of binding to DNA, an interaction that may lead to mutagenic and carcinogenic outcomes.^40^ Collectively, these pathophysiological processes, combined with effects from other PM_2.5_ constituents, can induce a cascade of cerebrovascular damage, including increased blood coagulability, plaque destabilization, accelerated atherosclerosis, and sustained vascular inflammation, all of which ultimately elevate the risk of stroke onset.^11,41,42^

The specific role of black carbon from traffic-related diesel combustion in stroke risk is supported by multiple lines of evidence. Short-term exposure has been shown to trigger the onset of large-artery atherosclerotic ischemic stroke,^43^ while long-term exposure has been associated with increased stroke incidence^44^ and subclinical precursors like increased carotid intima-media thickness.^45^ Notably, the large-scale ELAPSE study further confirmed that the adverse effect of black carbon on stroke incidence persists even at low concentrations, underscoring its potency as a cerebrovascular risk factor even in relatively clean environments.^46^ Beyond black carbon, organic matter has also emerged as a component of concern, particularly for impairing neurological recovery after stroke.^27^ Furthermore, the health effects of air pollution can be modified by meteorological conditions. A nationwide study in China demonstrated that heat waves interacted synergistically with PM_2.5_ constituents, especially secondary inorganic aerosols such as nitrate, sulfate, and ammonium, resulting in a greater-than-additive increase in stroke mortality.^47^

Stratified analyses revealed significantly stronger associations among older adults (≥ 65 years), a finding consistent with previous epidemiological evidence.^12,48,49^ This may be attributed to age-related physiological degradation, including cardiovascular aging and weakened immune function, which can amplify vulnerability to the cerebrovascular injury induced by PM_2.5_ constituents.^50^ Additionally, the progressive accumulation of cardiovascular risk factors over the life course, such as hypertension, diabetes, dyslipidemia, vascular stiffness, and endothelial dysfunction, may further exacerbate this susceptibility.^51^ Notably, many of these conditions are themselves influenced by exposure to PM_2.5_ constituents, suggesting a potential synergistic pathway in the development of stroke.^52–55^ Another notable finding was the elevated susceptibility among residents in southern China, a pattern consistent with existing evidence. A national screening program identified the highest carotid plaque prevalence in southwestern China (43.17%), a known precursor to stroke.^56^ Two recent studies on cardiovascular diseases and PM_2.5_ constituents also identified stronger adverse effects in southern populations.^29,57^ This north-south disparity likely arises from a combination of factors, including climatic conditions that promote secondary aerosol formation in the south, alongside differences in emission sources, pollution levels, and population characteristics.

In light of our results, targeted strategies are warranted to mitigate stroke risk from PM_2.5_ constituents. At the policy level, transitioning to a clean energy economy and phasing out industrial animal farming are critical steps.^58,59^ For vulnerable individuals, practical measures such as using air filtration systems, wearing protective masks outdoors, and scheduling activities around air quality forecasts are recommended.^60^ Furthermore, expanding urban green infrastructure can serve as an effective public health intervention to ameliorate the cardiovascular burden of air pollution.^61^

Our study possesses several key strengths. First, the prospective cohort study design with a nationwide population-based sample ensured the reliability of our conclusions, while a series of sensitivity analyses further confirms the stability and robustness of the observed associations. Second, environmental exposure data were derived from a model with high spatial and temporal resolution, thereby reducing exposure misclassification in comparison with the use of fixed-site monitoring stations alone. Third, the application of the QGC model enabled the examination of the individual contribution of each constituent, effectively addressing the collinearity inherent in mixtures of pollutants. However, several limitations in our study should be acknowledged. First, while our sample size is nationally representative and provided sufficient statistical power to detect robust associations (as evidenced by narrow confidence intervals), it is more modest than some studies utilizing large administrative databases. However, the major strength of CHARLS cohort lies in its rich, individual-level data on a wide array of potential confounders, enabling more rigorous confounding control. Second, exposure was estimated based on residential addresses without accounting for indoor pollution from activities like cooking or heating with solid fuels, which emit harmful particles including black carbon and organic matter. This may have introduced exposure misclassification. Future studies should incorporate automatic and portable monitoring to improve the accuracy of individual exposure assessment. Third, although we adjusted for a range of individual-level covariates, residual confounding remains possible. Additionally, both the QGC and WQS models provide fixed weights for mixture constituents without confidence intervals, limiting the ability to evaluate the statistical significance of these weights. Finally, the stroke cases in the CHARLS survey were not classified into subtypes. Therefore, our findings reflect the overall association between PM_2.5_ constituents and incident stroke, without distinguishing which subtype may be more severely affected. Well-designed, large-scale studies are warranted to investigate the relationships between specific PM_2.5_ constituents and stroke subtypes.

## 5 Conclusion

In summary, this prospective cohort study provides robust evidence linking long-term exposure to specific PM_2.5_ constituents with an increased risk of incident stroke among middle-aged and older adults in China. Our findings underscore the predominant role of carbonaceous components, particularly black carbon and organic matter. Given the rapidly aging population in China, these results hold significant implications for guiding future clinical preventive strategies, public health interventions, and targeted environmental policies to mitigate the stroke burden.

## Data Availability

The data supporting the findings of this study are accessible in the methods section of this article, and can be obtained from the corresponding author upon reasonable request.

https://www.ecmwf.int/en/forecasts/dataset/ecmwf-reanalysis-v5

http://tapdata.org.cn/

https://charls.pku.edu.cn/

## Acknowledgements

This study relied on publicly available data from CHARLS, TAP database, and the ERA5 reanalysis dataset. We thank all individuals and teams responsible for providing these essential resources.

## Supplemental Material

Tables S1–S5

Figures S1–S6

## Non-standard Abbreviations and Acronyms

DALY: disability-adjusted life year
PM2.5: fine particulate matter
CHARLS: China Health and Retirement Longitudinal Study
APPCAP: Air Pollution Prevention and Control Action Plan
TAP: Tracking Air Pollution in China
WRF-CMAQ: Weather Research and Forecasting-Community Multiscale Air Quality
ERA5: European Centre for Medium-Range Weather Forecasts Reanalysis v5
SBP: systolic blood pressure
DBP: diastolic blood pressure
BMI: body mass index
WC: waist circumference
SD: standard deviation
VIF: variance inflation factor
HR: hazard ratio
CI: confidence interval
IQR: interquartile range
RCS: restricted cubic spline
QGC: quantile-based g-computation
WQS: weighted quantile sum
AUC: area under the curve
PAH: polycyclic aromatic hydrocarbon
ROS: reactive oxygen species

